# Investigating the source of increased bipolar and major depressive disorder polygenic risk in multiplex schizophrenia families

**DOI:** 10.1101/2021.11.15.21266368

**Authors:** Mohammad Ahangari, Robert Kirkpatrick, Tan-Hoang Nguyen, Nathan Gillespie, Irish Schizophrenia Genomics Consortium, Kenneth S. Kendler, Silviu-Alin Bacanu, Bradley T. Webb, Brian C. Verrelli, Brien P. Riley

## Abstract

Psychotic and affective disorders often aggregate in the relatives of probands with schizophrenia (SCZ), and genetic studies show substantial genetic correlation among SCZ, bipolar disorder (BIP) and major depressive disorder (MDD). However, the nature of this genetic overlap in polygenic risk score (PRS) analyses of multiplex families has not been fully dissected. In the current study, we investigated the polygenic risk burden of BIP and MDD in a sample of 257 multiplex SCZ families (N=1,005) and population controls (N=2,205). Furthermore, due to the strong genetic correlation among SCZ, BIP, and MDD, we examined whether increased BIP or MDD PRS in members of multiplex SCZ families can be attributed to latent genetic factors unique to BIP or MDD, or latent genetic factors that each of these two disorders share with SCZ. Our results indicate that members of multiplex SCZ families have an increased PRS for BIP and MDD, however, this observation is largely attributable to latent genetic factors that BIP or MDD share with SCZ, rather than latent genetic factors unique to them. These results provide new insight for cross-disorder PRS analyses of psychiatric disorders, by cautioning that for complete interpretation of observed cross-disorder PRS enrichment, we should account for genetic correlations across psychiatric disorders. Our findings further indicates that members of multiplex SCZ families may have an increased genetic vulnerability to both psychotic and affective disorders, and for full assessment of an individual’s genetic risk, familial backgrounds should be taken into consideration.

## Introduction

Psychotic and affective disorders have long been viewed as two separate axes of mental illness, but early practitioners of psychiatry like Emil Kraepelin and Eugen Bleuler observed that relatives of patients with schizophrenia (SCZ) have an increased rate of psychotic and affective disorders, many of which appeared to be milder versions of the symptoms observed in patients with SCZ (1). Some of the first family studies of SCZ conducted in the early 20^th^ century, confirmed that in addition to SCZ, a range of other psychiatric disorders also aggregate in the relatives of probands with SCZ (2). These findings were later solidified by the Danish Adoption Study of Schizophrenia, which showed that biological relatives of patients with SCZ were at an increased risk for SCZ and a spectrum of other psychiatric disorders (3,4).

The Irish Study of High-Density Schizophrenia Families (ISHDSF) (5,6) consists of 257 multiplex SCZ families with genotype data, ascertained to have two or more first-degree relatives meeting the Diagnostic and Statistical Manual of Mental Disorders (DSM-III-R) criteria for SCZ or poor-outcome schizoaffective disorder (Supplementary Table 1). In line with previous epidemiological observations in the relatives of probands with SCZ (5,7,8), a broad spectrum of other psychiatric diagnoses, including psychotic, affective, personality, and substance use disorders, are also present in the ISHDSF sample (6,9–12), with previous polygenic risk score profiling in the ISHDSF sample showing that all family members, including the unaffected individuals, have an increased burden of SCZ common genetic risk variation compared to population controls (13).

Results from the Psychiatric Genomics Consortium (PGC) Cross-Disorder Group (14,15), and other cross-disorder analyses of psychiatric disorders (16,17) (Supplementary Figure 4), have provided robust, replicable evidence for strong genetic correlation (r_G_) between SCZ and bipolar disorder (BIP), and to a lesser degree between SCZ and major depressive disorder (MDD). The genetic correlation between SCZ and BIP is estimated to be r_G_ = 0.68, and of the 64 genome-wide significant loci associated with BIP, 10 have previously reached genome-wide significance level in SCZ (18). SCZ and MDD also have a significant positive genetic correlation estimated to be r_G_ = 0.35, and in addition to substantial overlap among genes identified to be involved in the genetic architecture of MDD and SCZ, 6 of the 44 loci associated with MDD, are also associated with SCZ (19). BIP and MDD also have significant positive genetic correlation estimated to be r_G_ = 0.44 (18).

The high baseline risk of SCZ observed across all diagnostic categories of ISHDSF sample, coupled with the evidence for cross-trait genetic correlations among SCZ, BIP, and MDD, and the presence of a wide spectrum of psychiatric disorders that aggregate in ISHDSF sample, raises two important questions. First, do members of multiplex SCZ families have an increased genetic risk for BIP and MDD? We constructed univariate BIP and MDD polygenic risk scores (PRS) to address the first question. Second, is the increased BIP or MDD genetic risk in multiplex SCZ families attributable to underlying latent genetic factors that BIP or MDD share with SCZ, or are they attributable to underlying latent genetic factors that are unique to BIP or MDD (hence, not shared with SCZ)? We used genomic structural equation modelling (genomicSEM) (20) and GWAS-by-subtraction (21) to perform a GWAS of latent, unmeasured factors underlying BIP or MDD to answer the second question. GWAS-by-subtraction is a newly developed method that facilitates the study and interpretation of residual values from genomicSEM models and provides insight into previously unmeasured latent genetic factors that are unique to individual phenotypes (21).

In addressing these two questions, we attempt to investigate the common risk variation burden of BIP and MDD, two co-morbid psychiatric disorders with SCZ, in families with multiply affected individuals with SCZ and related psychiatric disorders. Furthermore, we shed light on the complexity of cross-disorder PRS analyses of psychiatric disorders in multiplex families, by taking into consideration the underlying genetic correlation among SCZ, BIP, and MDD.

## Materials and Methods

### Sample Description

#### Irish Study of High-Density Schizophrenia Families (ISHDSF)

Fieldwork for the ISHDSF sample was carried out between 1987 and 1992, with probands ascertained from public psychiatric hospitals in the Republic of Ireland and Northern Ireland with approval from local ethics committees (6). Selection criteria were two or more first-degree relatives meeting DSM-III-R criteria for SCZ or poor-outcome schizoaffective disorder (PO-SAD), with all four grandparents being born in either Ireland or the United Kingdom. Relatives of the probands suspected of psychotic illness were interviewed by trained psychiatrists, and trained social workers interviewed other relatives of the probands. To avoid bias and detect possible mistakes in diagnosis, independent review of all diagnostic information was made blind to family assignments by two trained psychiatrists, with each psychiatrist making up to 3 best estimate DSM-III-R diagnoses, with high agreement (weighted k= 0.94 +-0.05).

The concentric diagnostic schema of the ISHDSF (Supplementary Figure 21), ranked by the degree to which they reflect the core vs periphery of the psychosis spectrum, includes 4 case definitions in the families as follows: *narrow* (SCZ, PO-SAD and simple SCZ), *intermediate* (adding schizotypal personality, schizophreniform, and delusional disorders, psychosis not otherwise specified (NOS), and good-outcome schizoaffective disorder, diagnoses that robustly and replicably aggregate in the relatives of SCZ probands), *broad* (adding psychotic affective illness, paranoid, avoidant and schizoid personality disorders and other disorders that significantly aggregated in relatives of SCZ probands in the Roscommon Family(10)), and *very broad* (adding any other psychiatric illnesses in the families). The ISHDSF sample also includes *unaffected* family members with no diagnosis of any psychiatric illness. Details of the ISHDSF sample are described elsewhere (6,22).

#### Population controls and replication singleton cases from Irish Schizophrenia Genomics Consortium

The Irish Schizophrenia Genomic Consortium (ISGC) sample was assembled for a GWAS of SCZ in Ireland. Details of recruitment, screening, and quality control (QC) are described elsewhere (23). Briefly, controls from the Irish Biobank used in ISGC were blood donors from the Irish Blood Transfusion Service recruited in the Republic of Ireland. Individuals reported all four grandparents born in either Ireland or the United Kingdom, with no reported history of psychotic illness. Due to the relatively low lifetime prevalence of SCZ in the general population (∼1%), misclassification of controls should have a minimum impact on power (24). Singleton SCZ cases used as a replication in this study were recruited through community mental health service and inpatient units in the republic of Ireland or Northern Ireland following protocols with local ethics approval. All participants were interviewed using a structured clinical interview for DSM-III-R or DSM-IV, were over 18 years of age and reported all four grandparents born in Ireland or the United Kingdom. Cases were also screened to exclude substance-induced psychotic disorder or psychosis due to a general medical condition. Detailed description for sample recruitment is provided in detail elsewhere (13,25).

### Genotyping and QC

Genotyping was carried out on 3 different arrays. 830 individuals from ISHDSF sample were genotyped on the Illumina 610-Quad Array. An additional 175 ISHDSF individuals, which either were not included in the Illumina Array study or did not pass QC, were later genotyped on the Infinium psychArray V.1.13 Array (the psychArray). For the ISGC sample, 1,730 population controls and 1,627 singleton cases were genotyped using the Affymetrix V.6.0 Array. An additional 475 population controls and 487 singleton cases that either did not pass the QC or were not included in the Affymetrix Array study were later genotyped on the psychArray along with the additional ISHDSF individuals. The same QC protocols were applied to all three datasets. In brief, exclusion criteria for samples were a call rate of <95%, more than one Mendelian error in the ISHDSF sample, and difference between reported and genotypic sex. Exclusion criteria for SNPs were MAF <1%, call rate <98%, and p<0.0001 for deviation from Hardy-Weinberg expectation. The final ISHDSF sample includes 1,005 individuals from 257 pedigrees whose SNP data from the Illumina Array and the psychArray passed all the QC filters. The final ISGC sample includes 2,205 controls and 2,114 singleton cases whose SNP data from the Affymetrix and the psychArray passed all the QC filters (Supplementary Table 2).

### Imputation

Genotypes passing QC were phased using Eagle V.2.4 (26) and imputed to the Haplotype Reference Consortium (HRC) reference panel (27) on the Michigan Imputation Server using Minimac4 (28). The HRC panel includes 64,975 samples from 20 different studies that are predominantly of European ancestry, making the HRC suitable for imputation of our homogenous sample from Ireland. Each of the three genotype sets described in the previous step was imputed separately, and the imputed genotype probabilities were downloaded in VCF format from the Michigan Imputation Server. Genotype dosages were extracted and used for all downstream analyses. As part of the post-imputation QC, variants with MAF <1% and r^2^ score of < 0.3 (29) were excluded. After imputation and all QC measurements, 9,298,012 SNPs on the Illumina Array, 11,080,279 SNPs on the Affymetrix Array, and 11,081,999 SNPs on the psychArray, remained for analysis. Of these three sets of SNPs, 9,008,825 SNPs were shared across all three imputed arrays and were used for downstream analyses. Description of QC steps for the imputation are described in the Supplementary Materials and Supplementary Figures 1-3.

### Summary statistics acquisition

We made use of the publicly available summary statistics data. PGC3-SCZ (67,390 cases and 94,015 controls) (30) and PGC3-BIP (41,917 cases and 371,549 controls) (18) summary statistics were downloaded from the PGC website. We used the 2019 MDD GWAS summary statistics from a meta-analysis of the PGC2-MDD and UK Biobank (excluding 23andMe), containing 170,756 cases and 329,443 controls (31). Low-density lipoprotein (LDL) summary statistics (N=87,048) (32) used as a negative control in this study was obtained from the ENGAGE Consortium website.

### GWAS-by-subtraction

We investigated whether increased BIP and MDD PRS in multiplex families can be attributed to underlying latent genetic factors that are unique to BIP or MDD (not shared with SCZ), or underlying latent genetic factors that BIP or MDD share with SCZ. We performed GWAS-by-subtraction within the genomicSEM framework by analyzing summary statistics data for SCZ (N=161,405), BIP (N=413,466), and MDD (N=500,199), by regressing SCZ and BIP or SCZ and MDD summary statistics on two latent variables that we called *SCZ* factor and *nonSCZ* factor. Subsequently, we regressed *SCZ* factor and *nonSCZ* factor on each SNP from the summary statistics, which allowed for two paths of association with BIP or MDD for each SNP: 1) a first path that is fully mediated by *SCZ* factor, and 2) a second path that is fully independent of *SCZ* factor, called *nonSCZ* factor (Supplementary Figure 5). Detailed description of GWAS-by-subtraction framework is provided in the original publication by Demange et al (21) and description of the current analysis, path estimates as well as the formula used to estimate the effective sample size for GWAS-by-subtraction analyses are provided in the Supplementary Materials and Supplementary Table 4.

### Estimation of SNP based heritability

We used LDSC (33) to estimate the SNP-based heritability of the genomicSEM GWAS results and partition the heritability into functional categories and cell types. Since genomicSEM results rely on latent factors, it is not possible to estimate the heritability on liability scale, therefore, all the SNP-based heritability estimates are reported on the observed scale.

### Construction of Polygenic Risk Score

Summary statistics for BIP (N=413,466), MDD (N=500,199, *SCZ* factor underlying BIP (N_*eff*_=149,460), *nonSCZ* factor underlying BIP (N_*eff*_ = 312,118), *SCZ* factor underlying MDD (N_*eff*_ = 149,464), and *nonSCZ* factor underlying MDD (N_*eff*_ = 461,356) were first QC’d by excluding variants with MAF < 1% or imputation quality score of < 0.9, and removing strand ambiguous and in/del polymorphisms. We then constructed PRS using a Bayesian regression framework by placing a continuous shrinkage prior on SNP effect sizes using PRS-CS (34). PRS-CS uses LD information from an external reference panel (the 1000 Genomes European Phase 3 European sample here) (35), to estimate the posterior effect sizes for each SNP. Although *p*-value thresholding and clumping method (P-T) have been traditionally used for PRS construction (36), PRS-CS has shown substantial improvement in predictive power over P-T (37). Similar to LDSC, PRS-CS limits the SNPs for PRS construction to around 1.2 million high-quality variants from the HapMap3 variants which provides ∼ 500 SNPs per LD block which substantially reduces memory and computational costs.

To show the specificity of the PRS constructed in our analysis, an additional PRS for low-density lipoprotein (LDL, N=87,048) from the ENGAGE Consortium (32) was also constructed using the same protocol described above. Genetic correlation estimates show that there is no significant correlation between LDL and psychiatric disorders, making LDL an appropriate phenotype as a negative control for this analysis (38,39).

### Genomic Relationship Matrix, Principal Component and Statistical Analyses

To account for the high degree of relatedness among individuals, analyses were carried out using a mixed-effects logistic regression model fitted by maximum likelihood by Nelder-Mead optimization using the GMMAT package (40) in R (41) The family structure was modelled as a random effect, with genomic relationship matrix (GRM) calculated using LDAK with LD correction parameters suited for families (42). In order to account for batch effects due to genotyping carried out on different arrays, we also included platform as a covariate. Principal component analysis (PCA) shows that all individuals in the sample are of European ancestry (Supplementary Figure 6-8). But to account for fine-scale structure within the Irish population (Supplementary Figure 9), the top 10 principal components (PC) were also included as covariates in the analysis. The final mixed regression model included GRM as a random effect covariate, with top 10 PCs, platform, and sex as fixed effect covariates. The final results were adjusted for multiple-comparison using the Holm method. In order to generate comparable odds ratios, all PRS underwent Z-score normalization.

## Results

### Multiplex SCZ families have an increased PRS for BIP and MDD

Table 1 presents the results from logistic mixed models per comparison group. As expected, LDL PRS used as the negative control in this study, shows no significant increase in members of multiplex SCZ families compared to population controls. Univariate BIP PRS was significantly higher in all diagnostic categories of multiplex SCZ families compared to population controls, except the unaffected individuals (only nominally significant). The highest odds ratio (OR) was observed in the *broad* category (OR = 2.21, 95% CI = 1.57-2.89), which includes 17 of the 21 BIP diagnosis in the ISHDSF. (Table 1). With the exception of the unaffected individuals in the families, MDD PRS was also significantly higher in all diagnostic categories compared to population controls, with the highest OR observed in the *very broad* category (OR = 1.52, 95% CI = 1.20-1.75), which includes 80 of 102 MDD diagnosis in the ISHDSF, excluding MDD cases with psychotic features (Supplementary Table 1).

**Table 1:** Comparison of the PRS in all diagnostic categories of ISHDSF from mixed-effects logistic regression models. All the *p*-values are one-sided and results are following the hypothesis that diagnostic categories in the families have higher PRS than population controls. *P*-value column represents the significance level before multiple testing corrections. Holm-adjusted *P*-value column represents the *p*-values adjusted for multiple testing correction using the Holm method OR and 95% CI are provided for effect sizes. Singleton cases versus population controls comparison follows the hypothesis that singleton cases have higher PRS than population controls.

| PRS | OR | 95% CI | P-Value | Holm-adjusted |
| --- | --- | --- | --- | --- |
| **Narrow vs Population Controls** |  |  |  |  |
| BIP | 1.93 | 1.69-2.20 | 7.21E-23 | 2.02E-21 |
| MDD | 1.35 | 1.98-1.53 | 1.45E-06 | 2.61E-05 |
| SCZ Factor in BIP | 6.11 | 5.41-7.02 | 6.53E-69 | 2.48E-87 |
| nonSCZ Factor in BIP | 0.97 | 0.51-1.24 | 0.3 | 1 |
| SCZ Factor in MDD | 6.06 | 5.25-6.99 | 9.40E-60 | 3.67E-88 |
| nonSCZ Factor in MDD | 0.95 | 0.48-1.20 | 0.37 | 1 |
| LDL | 0.98 | 0.88-1.11 | 0.82 | 1 |
| **Intermediate vs Population Controls** |  |  |  |  |
| BIP | 1.84 | 1.48-2.29 | 5.85E-08 | 1.17E-06 |
| MDD | 1.43 | 1.17-1.75 | 5.72E-04 | 8.00E-03 |
| SCZ Factor in BIP | 5.76 | 4.72-7.22 | 7.63E-32 | 2.48E-30 |
| nonSCZ Factor in BIP | 0.96 | 0.54-1.19 | 0.16 | 1 |
| SCZ Factor in MDD | 5.78 | 4.70-7.22 | 7.50E-32 | 2.48E-30 |
| nonSCZ Factor in MDD | 0.95 | 0.654-1.16 | 0.11 | 1 |
| LDL | 0.85 | 0.70-1.04 | 0.13 | 1 |
| **Broad vs Population Controls** |  |  |  |  |
| BIP | 2.21 | 1.57-2.89 | 7.32E-07 | 1.39E-05 |
| MDD | 1.41 | 1.05-1.90 | 2.14E-02 | 2.58E-01 |
| SCZ Factor in BIP | 4.39 | 2.95-5.64 | 6.61E-15 | 1.59E-13 |
| nonSCZ Factor in BIP | 0.99 | 0.517-1.26 | 0.27 | 1 |
| SCZ Factor in MDD | 4.22 | 2.87-5.83 | 4.49E-15 | 1.12E-13 |
| nonSCZ Factor in MDD | 0.98 | 0.63-1.11 | 0.23 | 1 |
| LDL | 0.88 | 0.62-1.08 | 0.16 | 1 |
| **Very Broad vs Population Controls** |  |  |  |  |
| BIP | 1.50 | 1.23-1.83 | 5.60E-05 | 8.96E-04 |
| MDD | 1.52 | 1.20-1.75 | 1.20E-04 | 1.80E-03 |
| SCZ Factor in BIP | 3.89 | 2.90-5.01 | 3.64E-26 | 1.09E-24 |
| nonSCZ Factor in BIP | 0.97 | 0.60-1.20 | 0.43 | 1 |
| SCZ Factor in MDD | 3.85 | 2.88-4.92 | 7.20E-25 | 2.09E-23 |
| nonSCZ Factor in MDD | 0.94 | 0.66-1.18 | 0.51 | 1 |
| LDL | 1.01 | 0.84-1.22 | 0.87 | 1 |
| **Unaffected vs Population Controls** |  |  |  |  |
| BIP | 1.21 | 1.03-1.40 | 0.003 | 0.06 |
| MDD | 1.1 | 0.94-1.27 | 0.23 | 1 |
| SCZ Factor in BIP | 3.60 | 2.89-4.70 | 1.36E-34 | 4.76E-33 |
| nonSCZ Factor in BIP | 0.95 | 0.54-1.21 | 0.33 | 1 |
| SCZ Factor in MDD | 3.44 | 2.80-4.68 | 1.16E-33 | 3.94E-32 |
| nonSCZ Factor in MDD | 0.92 | 0.50-1.18 | 0.55 | 1 |
| LDL | 0.98 | 0.84-1.14 | 0.85 | 1 |
| **Singleton Cases vs Population Controls** |  |  |  |  |
| BIP | 1.75 | 1.63-1.88 | 1.59E-52 | 5.88E-51 |
| MDD | 1.35 | 1.26-1.44 | 2.73E-18 | 7.37E-17 |
| SCZ Factor in BIP | 5.35 | 4.55-6.19 | 1.30E-22 | 5.33E-20 |
| nonSCZ Factor in BIP | 0.98 | 0.60-1.25 | 0.22 | 1 |
| SCZ Factor in MDD | 5.11 | 4.40-5.91 | 4.08E-22 | 1.71E-20 |
| nonSCZ Factor in MDD | 0.97 | 0.54-1.21 | 0.18 | 1 |
| LDL | 0.99 | 0.93-1.06 | 0.92 | 1 |

#### Increased BIP and MDD PRS in multiplex SCZ families are due to underlying *SCZ* factors

Next, we investigated whether the increased BIP and MDD polygenic risks in members of multiplex SCZ families are attributable to latent genetic factors that BIP or MDD share with SCZ, or to latent genetic factors that are unique to each disorder. (Supplementary Figure 4 for path diagram of the Cholesky decomposition). Using LDSC, we estimate the SNP-based heritability for *SCZ* factor underlying BIP 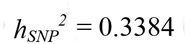 (se = 0.0112), *nonSCZ* factor underlying BIP 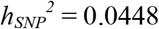 (se = 0.0023), and SCZ factor underlying MDD 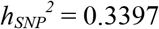 (se = 0.0116), *nonSCZ* factor underlying MDD 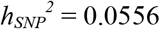 (se = 0.0022) (Supplementary Table 3). Manhattan and Q-Q plots for GWAS-by-subtraction models are represented in Figure 1 and 2, with full GWAS-by-subtraction results reported in in Supplementary Figures 6-11 and Supplementary Tables 3,4 and 13-18.

**Figure 1.**
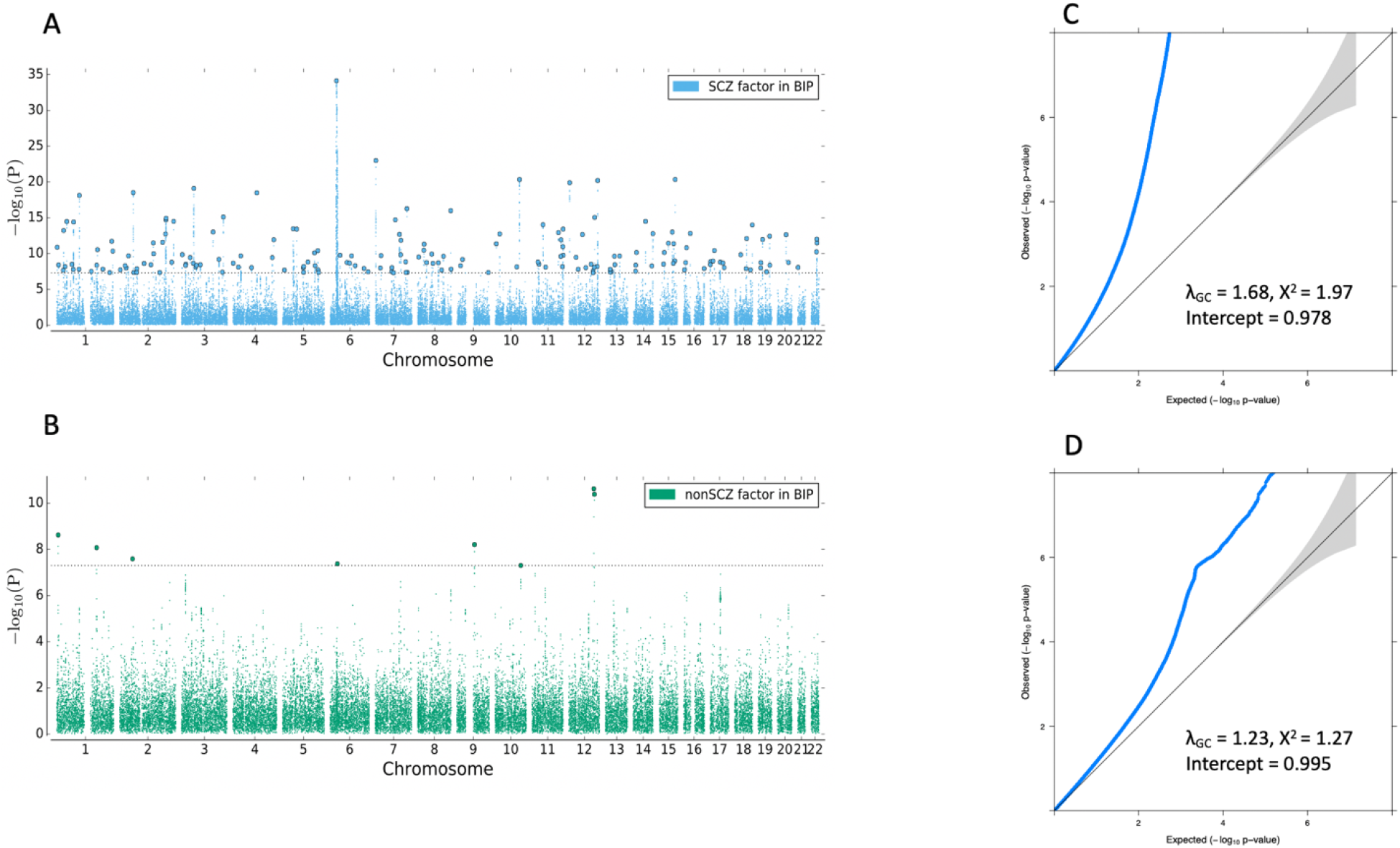
Manhattan and quantile-quantile plots for *SCZ* factor and *nonSCZ* factor in BIP. **A**. Manhattan plot corresponding to the SNP effects for SCZ factor in BIP. **B**. Manhattan plot corresponding to the SNP effects for nonSCZ factor in BIP. The x-axis in **A** and **B** corresponds to the chromosome and the y-axis shows the *p*-value on the -log_10_ scale. The dotted line denotes genome-wide significance level of 5×10^−8^. Lead SNPs are marked in bold. **C**. Quantile-quantile plot for *SCZ* factor in BIP. **D**. Quantile-quantile plot for *nonSCZ* factor in BIP. The x-axis in **C** and **D** refers to expected *p*-value while the y-axis refers to the observed *p*-value.

**Figure 2.**
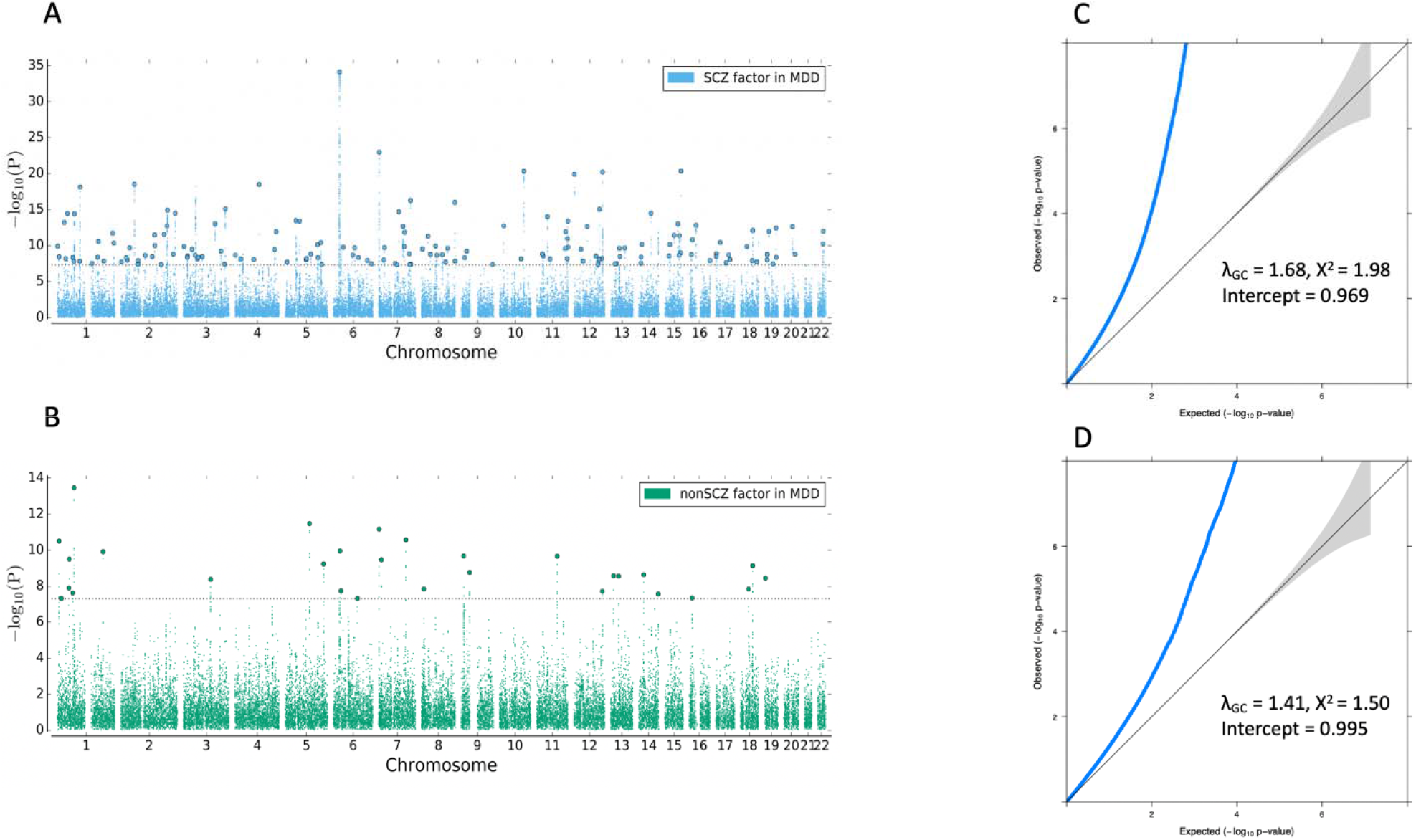
Manhattan and quantile-quantile plots for *SCZ* factor and *nonSCZ* factor in MDD. **A**. Manhattan plot corresponding to the SNP effects for SCZ factor in MDD. **B**. Manhattan plot corresponding to the SNP effects for nonSCZ factor in MDD. The x-axis in **A** and **B** corresponds to the chromosome and the y-axis shows the *p*-value on the -log_10_ scale. The dotted line denotes genome-wide significance level of 5×10^−8^. Lead SNPs are marked in bold. **C**. Quantile-quantile plot for *SCZ* factor in MDD. **D**. Quantile-quantile plot for *nonSCZ* factor in MDD. The x-axis in **C** and **D** refers to expected *p*-value while the y-axis refers to the observed *p*-value.

The PRS constructed for *SCZ* factor underlying BIP and *SCZ* factor underlying MDD were significantly higher in all diagnostic categories of multiplex SCZ families compared to population controls, with the highest OR observed in the *narrow* category (*SCZ factor* in BIP OR = 6.11, 95% CI = 5.41-7.02; *SCZ factor* in MDD OR = 6.06, 95% CI = 5.25-6.99) (Table 1, Figures 3,4). PRS constructed from *nonSCZ* factor in BIP and *nonSCZ* factor in MDD showed no significant increase in members of multiplex SCZ families compared to population controls (Table 1, Figures 3,4), indicating that the increased polygenic risk burden of BIP and MDD in multiplex SCZ families is likely to be fully attributable to the underlying latent genetic factor that BIP or MDD share in common with SCZ, as opposed to latent genetic factors or liabilities that are unique to BIP or MDD.

**Figure 3.**
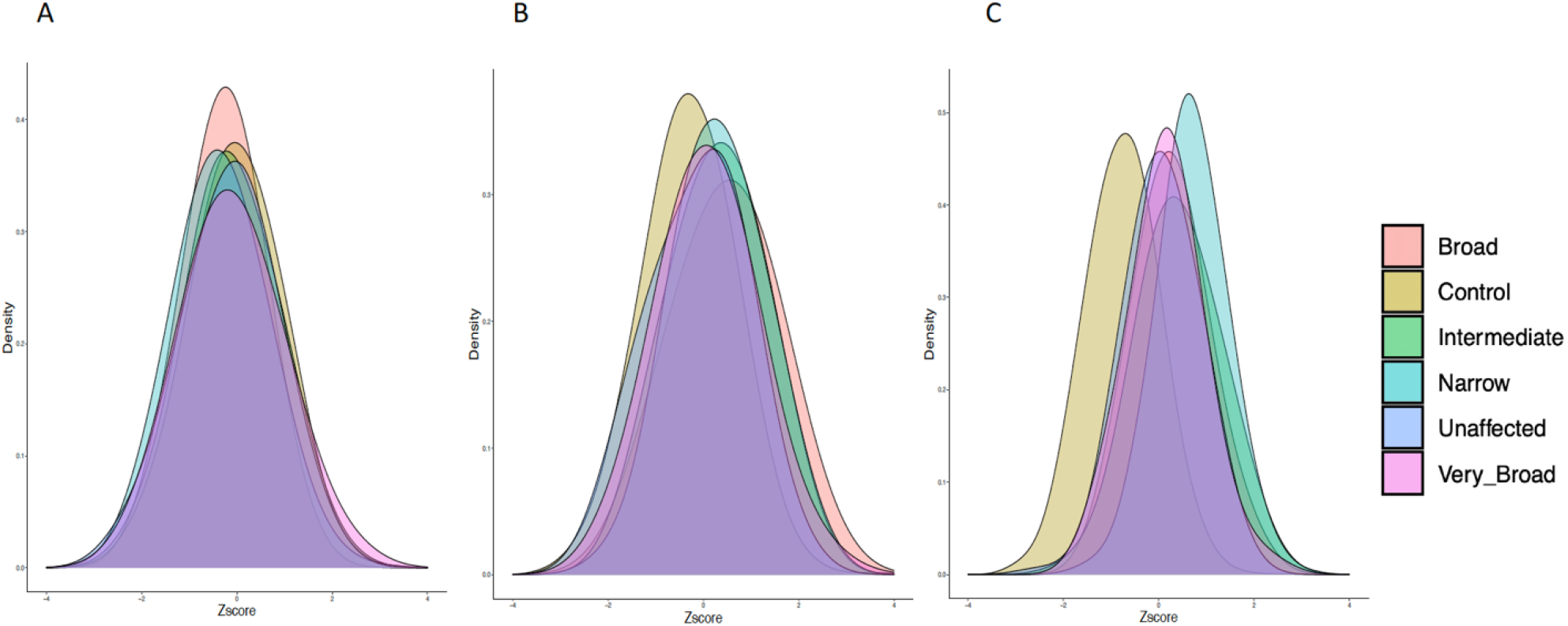
Density plots visualizing the distribution of BIP PRS in the ISHDSF sample. All PRS are Z-score standardized with mean of 0 and standard deviation of 1. **A**. Distribution of *nonSCZ* factor in BIP PRS. **B**. Distribution of univariate BIP PRS. **C**. Distribution of *SCZ* factor in BIP PRS. No significant difference is observed for the *nonSCZ* factor PRS between different diagnostic categories in ISDHSF versus to population controls, whereas *SCZ* factor PRS is significantly higher in all categories compared to population controls. Each color represents one of the diagnostic categories in ISHDSF sample on the psychosis spectrum (Supplementary Table 1 and Supplementary Figure 16)

**Figure 4.**
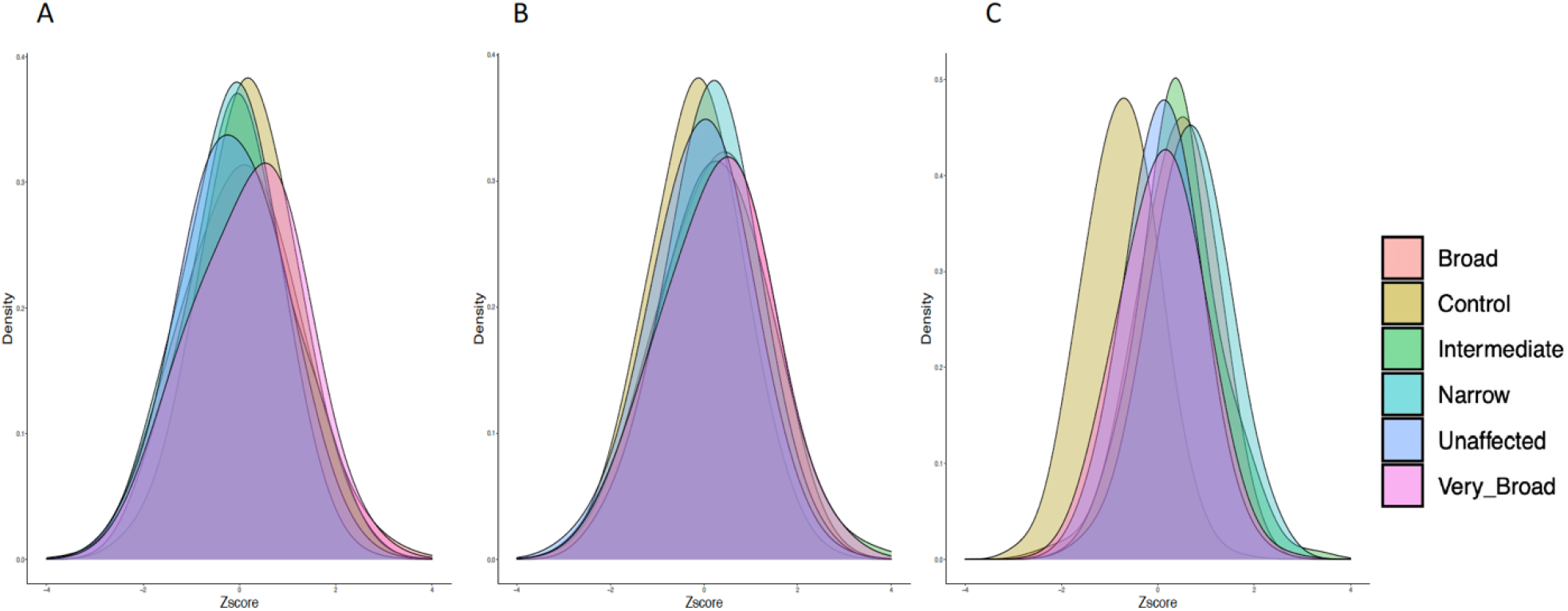
Density plots visualizing the distribution of MDD PRS in the ISHDSF sample. All PRS are Z-score standardized with mean of 0 and standard deviation of 1. **A**. Distribution of *nonSCZ* factor in MDD PRS. **B**. Distribution of univariate MDD PRS. **C**. Distribution of *SCZ* factor in MDD PRS. No significant difference is observed for the *nonSCZ* factor PRS between different diagnostic categories in ISDHSF versus to population controls, whereas *SCZ* factor PRS is significantly higher in all categories compared to population controls. Each color represents one of the diagnostic categories in ISHDSF sample on the psychosis spectrum (Supplementary Table 1 and Supplementary Figure 16)

### Replication

To demonstrate the generalizability of the results beyond multiplex families, we attempted a replication of the PRS comparison, in an independent sample of ancestry-matched singleton SCZ cases (N=2,224) from Ireland. The observed pattern of PRS enrichment in singleton SCZ cases is similar to the *narrow* category in multiplex SCZ families (which includes familial SCZ cases), showing the generalizability of these results in a cohort of non-related singleton SCZ cases from the sampe population. (Table 1).

## Discussion and Conclusions

Large-scale GWAS of SCZ, BIP, and MDD (18,19,30,31,43–45), have shown that many common risk variants with small effect sizes contribute to disease risk in psychiatric disorders, with cross-disorder analyses of psychiatric disorders also providing consistent evidence that SCZ, BIP, and MDD share substantial genetic risk at common variation level (16,17,46). In light of these observations, we investigated the common genetic risk variation burden of BIP, MDD in multiplex SCZ families, and further disentangled the observed increased polygenic risks into underlying latent genetic factors. Our results indicate that members of multiplex SCZ families have an increased PRS for BIP and MDD, with GWAS-by-subtraction analyses showing that this increased polygenic risk is likely to be entirely attributable to genetic risk factors that BIP or MDD share with SCZ. In addition, LDL PRS used as a negative control, showed no significant difference between members of multiplex SCZ families and population controls, validiting the observed patterns of PRS enrichment for underlying genetic factors generated in this study. Furthermore, we replicated our findings in an independent, ancestry-matched sample of singleton SCZ cases, to show the generalizability of this observation in a cohort of singleton SCZ cases. Our results provide genetic evidence in support of previous epidemiological findings that shows an increased incidence of both psychotic and affective disorders in families with multiple SCZ cases (5,6,13,47). Therefore, individuals in multiplex families may be genetically vulnerable to a range of psychotic and affective disorders, and in order to properly assess an individual’s genetic risk for psychiatric disorders, familial backgrounds should be taken into consideration.

GenomicSEM framework results rely on latent factors that are inferred from the molecular data (20). Therefore, we attempted to further validate the GWAS-by-subtraction models by conducting comprehensive downstream follow-up analyses. We show that *SCZ* and *nonSCZ* factor GWAS results generated from GWAS-by-subtraction models show strong polygenic signals with no evidence for confounding (Figures 1,2), and partitioning the heritability of these results into functional categories (48) reveals that both underlying factors for BIP or MDD are enriched in similar functional categories (Supplementary Tables 5-8). Furthermore, we show that GWAS-by-subtraction results are also significantly enriched in CNS tissues relevant to psychiatric disorders (Supplementary Figure 14 and Supplementary Table 17), and MAGMA tissue expression profile analysis (49) using GTEx v8 (50) also shows that genes from GWAS-by-subtraction analyses were significantly enriched for expression in nearly all central nervous system (CNS) tissues (Supplementary Figures 10-13 supplementary Table 15), with cell-type stratified LDSC (51) analyses also showing significant enrichment of genes found in neurons for GWAS-by-subtraction results, with no significant enrichment in oligodendrocytes and astrocytes (Supplementary Figure 14 and Supplementary Table 16), further underlining the validity of these results.

We note that increased polygenic risk for psychiatric disorders has been observed in other family and pedigree studies (52–54), however, the scope of the current study differs from previous studies. A distinct feature of the ISHDSF sample, is the presence of a broad range of psychiatric disorders, which allows for proper cross-disorder PRS analysis. In addition, to our knowledge, the ISHDSF sample also has the largest sample size (N=1,005) among currently published multiplex family PRS studies, and is the first to dissect polygenic risks into underlying genetic factors. Andlauer et al. (52) analyzed multiplex BIP families (N=395) consisting of 166 BIP and 78 MDD cases, and showed that familial BIP cases and their unaffected relatives, had a higher PRS for SCZ and BIP compared to population controls. Szatkiewicz et al. (53) used a densely affected pedigree from Northern Sweden (N=418) and showed an increased SCZ PRS in affected members compared to unaffected members and population controls. De Jong et al. (54) also used a dense pedigree (N=300) with BIP and MDD cases and showed nominally significant BIP and SCZ PRS in affected members compared to unaffected members and population controls. Our results, combined with previous cross-disorder analyses of multiplex families referenced above, suggest that we are only just beginning to tease apart the complex interactions underlying psychiatric disorders. Thus, as current GWAS sample sizes continue to increase, we will continue to reveal how these underlying genetic factors, both independently and through their covariance, contribute to complex psychiatric disorders and their genetic architectures at common variation level.

The analyses presented in this study should be interpreted in the context of some limitations.

The ISHDSF sample was ascertained to have 2 or more probands with SCZ or poor-outcome schizoaffective disorder. Therefore, some diagnostic categories in the families (e.g the *broad* category), have a lower number of individuals, which may potentially bias some of the results due to lower power. We addressed this by repeating the PRS analysis by grouping the individuals in a concentric manner as described in the original ISHDSF publication (6). The concentric comparison versus population controls showed similar patterns of PRS enrichment as observed in the separate comparisons presented in the main text, indicating that lower numbers of individuals in some of the diagnostic categories is unlikely to be a source of bias (Supplementary Table 19). Given that environmental factors have not been assessed here, future analyses could integrate environmental influences unique to the families to further elucidate the role of environmental factors on the elevated polygenic risk for BIP and MDD in multiplex SCZ families.

In conclusion, in this study we showed that members of multiplex SCZ families have an increased polygenic risk for BIP and MDD compared to ancestry-matched population controls. However, this observation is likely to be entirely attributable to latent genetic factors shared between SCZ and BIP, or SCZ and MDD, rather than latent genetic factors unique to BIP or MDD. These findings provide new insight for cross-disorder PRS analyses in psychiatric disorders, by cautioning that for complete interpretation of observed cross-disorder PRS enrichment, we must account for genetic correlations across correlated psychiatric disorders, as failure to do so, may result in erroneous conclusions about not only independent factors that contribute to polygenic architecture of complex psychiatric disorders, but also the degree to which complex psychiatric disorders are unique or related to each other.

## Data Availability

Code Availability:
All the scripts used in this study will be made available upon publication.
We made use of various freely available software tools in this study:
Plink2: https://www.cog-genomics.org/plink/2.0/
GenomicSEM: https://github.com/GenomicSEM/GenomicSEM
PRS-CS: https://github.com/getian107/PRScs
MiXeR: https://github.com/precimed/mixer
LDSC: https://github.com/bulik/ldsc
GMMAT: https://github.com/hanchenphd/GMMAT
LDAK: http://dougspeed.com/ldak/
Data Availability:
GWAS summary statistics for SCZ, BIP, MDD, and LDL are publicly available.
PGC3-SCZ: https://www.med.unc.edu/pgc/download-results/
PGC3-BIP: https://www.med.unc.edu/pgc/download-results/
PGC2-MDD-UKB Meta-analysis: https://datashare.ed.ac.uk/handle/10283/3203
LDL: http://diagram-consortium.org/2015_ENGAGE_1KG/
GWAS-by-subtraction summary statistics generated in this study will be made publicly available upon publication.

## Code Availability

All the scripts used in this study will be made publicly available upon publication.

We made use of various freely available software tools in this study:

Plink2:https:https://www.cog-genomics.org/plink/2.0/

GenomicSEM: https://github.com/GenomicSEM/GenomicSEM

PRS-CS: https://github.com/getian107/PRScs

MiXeR: https://github.com/precimed/mixer

LDSC: https://github.com/bulik/ldsc

GMMAT: https://github.com/hanchenphd/GMMAT

LDAK: http://dougspeed.com/ldak/

## Data Availability

GWAS summary statistics for SCZ, BIP, MDD, and LDL are publicly available.

PGC3-SCZ: https://www.med.unc.edu/pgc/download-results/

PGC3-BIP: https://www.med.unc.edu/pgc/download-results/

PGC2-MDD-UKB Meta-analysis: https://datashare.ed.ac.uk/handle/10283/3203

LDL: http://diagram-consortium.org/2015_ENGAGE_1KG/

GWAS-by-subtraction summary statistics generated in this study will be made publicly available upon publication.

## Funding and Acknowledgements

MA, BCV, S-AB, KSK, BTW and BR were supported by R01-MH114593 (to Dr. Riley). Production of GWAS data for singleton cases and controls was supported by R01-MH083094 (to Dr. Riley) and Wellcome Trust Case Control Consortium 2 project (085475/B/08/Z and 085475/Z/08/Z), and the Wellcome Trust (072894/Z/03/Z, 090532/Z/09/Z and 075491/Z/04/B), and NIMH grant MH 41953 and Science Foundation Ireland (08/IN.1/B1916). Production of GWAS data for multiplex families was supported by R01-MH062276 and R01-MH068881 (to Dr. Riley). T-HN was supported by NARSAD Young Investigator Grant 28599.

This study was approved by Virginia Commonwealth University Office of Research Subject Protection, and all necessary patient/participant consent has been obtained and the appropriate institutional forms have been archived.

<u>Membership of the Irish Schizophrenia Genomics Consortium (ISGC):</u>

Brien P Riley^1^, Derek W Morris^2^, Colm T O’Dushlaine^3^, Paul Cormican^4^, Elaine M Kenny^3^, Brandon Wormley^1^, Gary Donohoe^2^, Emma Quinn^3^, Roisin Judge^3^, Kim Coleman^3^, Daniela Tropea^3^, Siobhan Roche^5^, Liz Cummings^3^, Eric Kelleher^3^, Patrick McKeon^5^, Ted Dinan^6^, Colm McDonald^2^, Kieran C Murphy^7^, Eadbhard O’Callaghan^8^, Francis A O’Neill^9^, John L Waddington^10^, Ken S Kendler^1^, Michael Gill^3^, Aiden Corvin^3^

1 Depts of Psychiatry and Human Genetics, Virginia Institute of Psychiatric and Behavioral Genetics, Virginia Commonwealth University, Richmond, VA, USA;

2 Centre for Neuroimaging and Cognitive Genomics (NICOG), School of Psychology, National University of Ireland Galway, Ireland; School of Natural Sciences, National University of Ireland Galway, Ireland;

^3^ Neuropsychiatric Genetics Research Group, Institute of Molecular Medicine, Trinity College Dublin, Dublin, Ireland;

^4^ Animal and Grassland Research and Innovation Centre, Teagasc, Grange, Dunsany, County Meath, Ireland;

^5^ St Patrick’s University Hospital, James St., Dublin, Ireland;

^6^ Department of Psychiatry, University College Cork, Cork, Ireland;

^7^ Department of Psychiatry, Royal College of Surgeons in Ireland, Dublin, Ireland;

^8^ DETECT Early Psychosis Service, Blackrock, Co. Dublin, Ireland;

^9^ Centre for Public Health, Institute of Clinical Sciences, Queen’s University Belfast, Belfast, UK;

^10^ Molecular and Cellular Therapeutics, Royal College of Surgeons in Ireland, Dublin, Ireland;

